# Guideline Adherence and Subjective Effects of a Mobile Clinical Decision Support System on Physicians’ Practice: A Nationwide Survey-Based Within-Subject Study

**DOI:** 10.1101/2025.11.26.25341081

**Authors:** Eduardo Cardoso de Moura, Dayanna Quintanilha Palmer, Julia Guedes Valentim do Nascimento, Michelle Marques dos Santos, Renata de Almeida Pedro

## Abstract

**Study Design:** Cross-sectional, within-subject observational study using a nationwide survey of physicians.

**Objective:** To evaluate whether recent use of a mobile Clinical Decision Support System (CDSS) is associated with physicians’ perceived clinical update, adherence to guideline-based recommendations, and confidence in decision-making across common clinical conditions.

**Methods:** Between March and May 2025, 1,055 Brazilian physicians—active users of the Afya Whitebook® mobile CDSS—completed two standardized clinical vignettes each, randomly drawn from eight prevalent diseases covering acute and chronic conditions. Exposure was defined as self-reported consultation of the CDSS within the previous 24 hours. Primary outcomes included perceived clinical update (Likert 1–5), guideline-concordant decision (binary), and confidence in decision-making (Likert 1–5). Fixed-effects regressions with clustered standard errors at the participant level were applied, controlling for clinical case type and individual heterogeneity.

**Results:** A total of 4,220 responses were analyzed (1,054 CDSS; 3,166 control). Recent CDSS use was associated with higher perceived update (β = 0.22; 95% CI 0.12–0.32; p < 0.001), greater likelihood of guideline-concordant answers (β = 0.22; 95% CI 0.05–0.39; p = 0.013), and higher confidence (β = 0.12; 95% CI 0.04–0.20; p = 0.003), with consistent effects across conditions and subgroups.

**Conclusion:** Brief, real-world exposure to a mobile CDSS yielded small but meaningful improvements in both cognitive and affective dimensions of clinical practice, extending beyond decision accuracy to perceived confidence and sense of update. These findings highlight CDSS as complementary tools that reinforce evidence-based care while fostering continuous learning and professional assurance in everyday clinical decision-making.

**Key messages:**

- **What is already known on this topic:** Clinical Decision Support Systems (CDSS) improve adherence to evidence-based guidelines and reduce medical errors. However, their immediate and subjective effects—such as physicians perceived update and confidence—remain poorly understood.
- **What this study adds:** This nationwide within-subject study provides empirical evidence that recent use of a mobile CDSS (Afya Whitebook®) is associated with modest but statistically significant improvements in perceived clinical update, decision accuracy, and confidence. These findings expand understanding of the proximal cognitive and affective effects of CDSS use in everyday medical practice.
- **How this study might affect research, practice or policy:** CDSS may contribute not only to evidence-aligned decisions but also to a more positive professional experience. These effects should inform digital health strategies aimed at improving care quality and clinician well-being.

## INTRODUCTION

Digital transformation has profoundly reshaped the healthcare landscape, influencing every level from data management to day-to-day clinical practice. Among the emerging innovations, Clinical Decision Support Systems (CDSS) have gained prominence for providing real-time, evidence-based information that supports safer, more accurate, and up-to-date medical decisions (1,2). These systems have become critical allies in an era of exponentially expanding medical knowledge and increasing demands for continuous professional development. In 2023 alone, more than 1.5 million new citations were added to PubMed, highlighting the challenge clinicians face in keeping pace with the accelerating flow of scientific information (3).

CDSS have the potential to enhance the quality of care, patient safety, and the overall efficiency of healthcare services by promoting data-driven, evidence-based decisions (4). Systematic reviews and meta-analyses indicate that such tools improve adherence to clinical guidelines and reduce medical errors, although their effects on provider performance and clinical outcomes remain heterogeneous and strongly dependent on contextual factors such as workflow integration and user engagement (5,6).

Mobile versions of these systems, enabling rapid access to clinical recommendations at the point of care, have further expanded their reach—particularly in outpatient and primary care settings. Studies show that these applications facilitate adherence to protocols, support diagnostic reasoning, and are generally well accepted by physicians and medical students(7,8). In settings with limited connectivity, the possibility of offline access has been identified as a critical factor for feasibility (9).

Despite these advances, important knowledge gaps persist regarding the mechanisms through which digital tools influence clinicians’ decision-making behaviors and subjective experiences. A systematic review by Agarwal et al. (2021), conducted to inform a World Health Organization (WHO) guideline on digital decision-support tools in primary care, emphasized that available evidence remains of low or very low certainty, with heterogeneous results concerning adherence to recommended practices and quality of care. Moreover, none of the included studies assessed intermediate or proximal outcomes—such as the time from presentation to appropriate management, user acceptability, or resource utilization—revealing limited understanding of how digital tools exert their influence on clinical decisions(10).

In addition, a recent qualitative synthesis highlighted that the use of digital health technologies can affect not only technical performance but also cognitive and emotional dimensions, such as confidence, perceived usefulness, and mental workload (11). However, quantitative investigations exploring these subjective effects in an objective and measurable manner remain scarce—particularly in Latin American contexts, where medical training, digital infrastructure, and working conditions are highly heterogeneous.

In this context, the present study investigates, on a national scale, the proximal impact of recent and verified use of the Afya Whitebook® on three central dimensions of clinical decision-making: perceived clinical update, adherence to guideline-based recommendations, and decision confidence. Prior evidence indicates that the subjective experiences of healthcare professionals remain underexplored (11) while CDSS have consistently been shown to promote greater adherence to clinical recommendations (6,12,13). Accordingly, we test three hypotheses: physicians who accessed the CDSS within the past 24 hours **[1]** report a higher perceived level of clinical update for related diseases, **[2]** are more likely to select guideline-concordant management options, and **[3]** report greater confidence in their clinical decisions compared with situations in which they did not access the content.

## METHODS

We pre-registered our hypotheses (https://aspredicted.org/dd52-4c23.pdf) and the study received ethics approval from the Instituto de Ensino Superior Presidente Tancredo de Almeida Neves Ethics Committee (Approval number: 7.543.478).

### Participants

Participants consisted of physicians with an active registration in the Regional Medical Council of Brazil and active users of Afya Whitebook® (a Clinical Decision Support System — CDSS). We selected the physicians who accessed specific content regarding eight different popular diseases (acute ischemic stroke, asthma, type 2 diabetes mellitus, acute infectious gastroenteritis, systemic arterial hypertension, ST-segment elevation myocardial infarction, urinary tract infection, and community-acquired pneumonia) in the last 24 hours. Therefore, we used access to the system to identify physicians that were likely to have read and updated their knowledge about the accessed disease. A total of 112,461 physicians were invited to participate. Of those, 6,558 opened the Informed Consent Form, and 1,055 agreed to participate and completed the survey. To confirm this assessment and properly assign participants to the CDSS condition (had consulted the CDSS for the disease under investigation) or the control condition, participants confirmed that they had accessed the content in the questionnaire in the last 24 hours.

The study offered no financial incentives, and participation was voluntary and unpaid. Baseline variables were collected to characterize the sample (gender, age, geolocation, formation degree, medical specialty, and years since graduation), allowing for representativeness assessment and adjustment for potential confounders in the analyses.

In Brazil, there were 575,930 physicians as of early 2024, according to the recent edition of Demografia Médica (14). In comparison, the Afya Whitebook® platform registered approximately 150,000 monthly active physician users in the same year, being one of the most used clinical decision-support systems in Brazil, designed to support evidence-based practice by providing rapid access to structured, topic-specific medical content such as disease summaries, diagnostic pathways, therapeutic protocols, pharmacological information, and clinical calculators. The platform is available on both mobile and desktop devices, enabling physicians to access guideline adherent recommendations in real time across various clinical settings. At the time of data collection, the platform consisted exclusively of an author-curated content library and did not incorporate generative artificial intelligence tools (15, 16).

### Procedure

Participants first completed demographic and clinical background questions. They then evaluated their update level regarding the eight possible diseases under study. Next, they were presented to two clinical cases with two closed questions each about next steps in medical approach, each question followed by a measure of confidence in the response given. One of the clinical cases presented was selected based on the disease the participant accessed in the CDSS in the last 24 hours, increasing the likelihood of attribution to the CDSS condition, and the other one was randomly assigned among the seven other diseases under investigation. After evaluating each clinical case, participants were asked if they had accessed content about the disease of the respective clinical case in the last 24 hours. This variable was used to classify each physician into either the CDSS condition or control condition for each case. For instance, a physician who confirmed having accessed the content of pneumonia in the last 24 hours was classified in the CDSS condition for pneumonia case. Each respondent could be in the CDSS condition for one case and/or control for the other. Randomizing case order and including two clinical questions for each case strengthened robustness and the generalizability of results.

### Sample and Case Design

A total of 1,055 physicians participated, yielding 2,110 vignette clinical cases evaluated, and, therefore, 4,220 questions answered (1,054 in CDSS condition and 3,166 in control condition). The cases and questions were developed and validated by an independent panel of three internal medicine professors under the instruction of developing clinical cases for the diseases under investigation with following questions regarding further decision making on practical medical approach to the cases. The same independent panel achieved a consensus for validation of clinical cases and questions. All materials were presented in Portuguese, with translations available in appendix A, including the answers that were accepted as correct for each clinical case.

### Measures

The outcomes (DV) were:

- Self-reported guideline update level (H1): Measured in the question level as the self-reported update level for the disease correspondent to the question under investigation in a 5-point Likert scale (1 = “Not updated at all” and 5 = “Totally updated”);
- Correct answer (H2): Measured in the question level as a dummy variable whether the selected answer (out of four options) was correct, in other words, it matches current guideline-based recommendations;
- Perceived confidence in decision (H3): Measured in the question level as a 5-point Likert scale (1 = “Very Insecure” and 5 “Very secure”).

The independent variables (IV) were measured as:

- CDSS/control condition: Measured in the case level as a dummy variable whether the participant confirmed having accessed the disease content in the CDSS platform in the last 24 hours;
- Clinical case: Categorical variable, indicating which out of the 8 possible clinical cases is under investigation. Used as a control variable;
- Covariates: We also measured baseline variables to characterize our sample: gender, age, geolocation, formation degree, medical specialty, and years since graduation.

### Statistical Analysis

We conducted regressions with clustered standard errors and fixed-effects at the participant level using StataNow/BE 18.5. Fixed-effects models add respondent-specific intercepts to focus exclusively on within-respondent variation across questions, thereby accounting for any individual characteristics that do not vary across questions within respondents.

To test hypothesis 1, we performed a linear regression using the “Self-reported guideline update level” as the dependent variable and exposure to decision-support content (“CDSS condition”) as the independent variable, including “Clinical case” as control.

To test hypothesis 2, we performed a logistic regression using “Correct answer” as the dependent variable and exposure to decision-support content (“CDSS condition”) as the independent variable, including “Clinical case” as control.

To test hypothesis 3, we performed a linear regression using “Perceived confidence in decision” as the dependent variable and exposure to decision-support content (“CDSS condition”) as the independent variable, including “Clinical case” as control.

## RESULTS

### Descriptive Statistics

The total sample consisted of 4,220 questions evaluated by physicians with diverse clinical backgrounds. Regarding self-reported access to CDSS in the last 24 hours for the disease under investigation, 25.0% (n = 1,054) of the evaluations fell into the CDSS condition—indicating that the respondent had confirmed accessing the CDSS to consult information about the disease under investigation in the last 24 hours—while 75.0% (n = 3,166) were classified in the control condition.

Regarding participants’ profile, 52.9% of participants were females (n = 558). Most participants held no specialty degree and had no specialization ongoing (53,9%, n = 569), with 25.4% (n = 268) being specialists, and 20.7% (n = 218) currently in a specialization program.The average age was 33.6 years, and average years since graduation was 5.4 years, revealing a relatively young sample of physicians. Regarding geolocation, 45.6% (n = 481) were from the Southeast region, 26.0% (n = 274) from the Northeast region, 15.6% (n = 165) from the South region, 6.5% (n = 69) from the Central-West region, and lastly 6.3% (n = 66) from the North region of Brazil.

Attribution to the CDSS condition was self-reported, and each respondent could be in the CDSS condition for one case of two questions and/or control condition for the other case, and therefore being included with four questions in total in the full sample. In table 1, we compare descriptive statistics of the question-participant dyad between the CDSS and control conditions. We found no relevant difference between the conditions (table 1).

**Table 1.** Descriptive statistics - question-participant dyads by condition.

|  | Control condition | CDSS condition | Test | p-value |
| --- | --- | --- | --- | --- |
| Gender, N (%) |  |  |  |  |
| Female | 1,644 (51.9) | 588 (55.8) | $\chi^2(1) = 4.73$ | 0.030 |
| Male | 1,522 (48.1) | 466 (44.2) |  |  |
| Medical degree (%) |  |  |  |  |
| No specialty completed or ongoing | 1,696 (53.6) | 580 (55.0) | $\chi^2(2) = 1.00$ | 0.607 |
| First specialization ongoing | 654 (20.7) | 218 (20.7) |  |  |
| At least one specialty concluded | 816 (25.8) | 256 (24.3) |  |  |
| Brazilian Macroregion Location, N (%) |  |  |  |  |
| Southeast | 1444 (45.6) | 480 (45.5) | $\chi^2(4) = 5.03$ | 0.283 |
| Norhteast | 844 (26.7) | 252 (23.9) |  |  |
| South | 482 (15.2) | 178 (16.9) |  |  |
| Central-West | 206 (6.5) | 70 (6.6) |  |  |
| North | 190 (6.0) | 74 (7.0) |  |  |
| Age, mean (SD), y | 33.7 (8.9) | 33.3 (8.9) | $t(4218) = 1.36$ | 0.173 |
| Years since medical degree (SD), y | 5.4 (7.3) | 5.3 (7.4) | $t(4218) = 0.39$ | 0.697 |

In table 2, we report the balance of questions answered for each case under both conditions, stating no relevant difference between the groups. We also included the clinical case as a control variable in our models.

**Table 2.** Questions evaluated by clinical case by condition.

| Clinical Case, N (%) | Control condition | CDSS condition | Total |
| --- | --- | --- | --- |
| Asthma | 436 (13.8) | 162 (15.4) | 598 (14.2) |
| Ischemic stroke | 414 (13.1) | 122 (11.6) | 536 (12.7) |
| Diabetes mellitus type 2 | 346 (10.9) | 96 (9.1) | 442 (10.5) |
| Infectious gastroenteritis | 400 (12.6) | 84 (8.0) | 484 (11.5) |
| Hypertension | 318 (10.0) | 118 (11.2) | 436 (10.3) |
| Acute myocardial infarction | 326 (10.3) | 106 (10.1) | 432 (10.2) |
| Urinary tract infection | 488 (15.4) | 188 (17.8) | 676 (16.0) |
| Pneumonia | 438 (13.8) | 178 (16.9) | 616 (14.6) |
| Total | 3166 | 1054 | 4220 |

### Hypothesis Testing

Consistent with H1, Model 1 (table 3) demonstrated that physicians who reported having recently accessed the CDSS to consult information about a specific disease perceived themselves as more updated regarding that condition compared with diseases they had not accessed (β = 0.22; 95% CI 0.12–0.32; p < 0.001). Supporting H2, Model 2 (table 3) showed that physicians in the CDSS condition were significantly more likely to select the correct answer to the clinical questions than when in the control condition (β = 0.22; 95% CI 0.05– 0.39; p = 0.013). Finally, aligned with H3, Model 3 (table 3) indicated that the use of the CDSS also increased physicians’ confidence in their selected answer compared with the control condition (β = 0.12; 95% CI 0.04–0.20; p = 0.003).

**Table 3.**
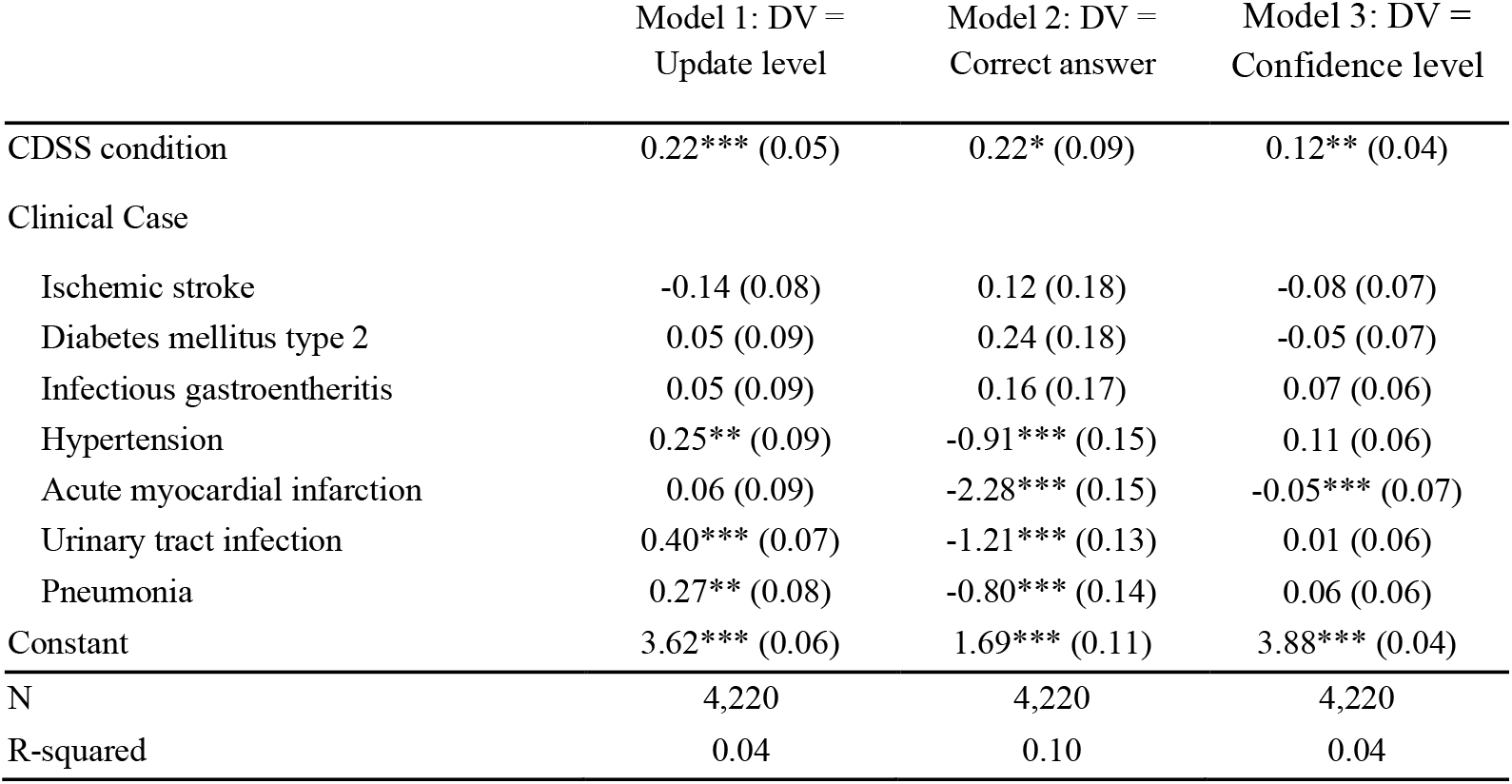
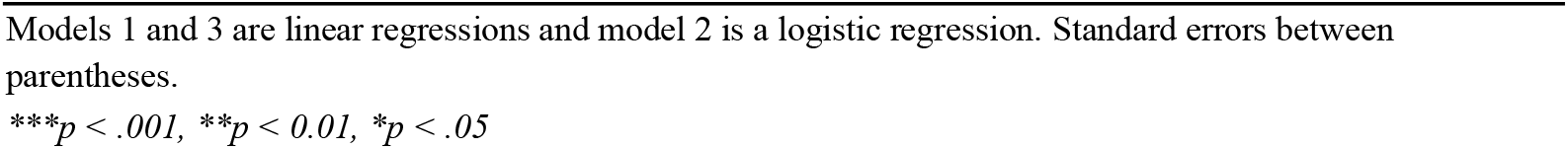
Regression models with clustered standard errors.

|  | Model 1: DV =<br>Update level | Model 2: DV =<br>Correct answer | Model 3: DV =<br>Confidence level |
| --- | --- | --- | --- |
| CDSS condition | 0.22*** (0.05) | 0.22* (0.09) | 0.12** (0.04) |
| Clinical Case |  |  |  |
| Ischemic stroke | -0.14 (0.08) | 0.12 (0.18) | -0.08 (0.07) |
| Diabetes mellitus type 2 | 0.05 (0.09) | 0.24 (0.18) | -0.05 (0.07) |
| Infectious gastroenteritis | 0.05 (0.09) | 0.16 (0.17) | 0.07 (0.06) |
| Hypertension | 0.25** (0.09) | -0.91*** (0.15) | 0.11 (0.06) |
| Acute myocardial infarction | 0.06 (0.09) | -2.28*** (0.15) | -0.05*** (0.07) |
| Urinary tract infection | 0.40*** (0.07) | -1.21*** (0.13) | 0.01 (0.06) |
| Pneumonia | 0.27** (0.08) | -0.80*** (0.14) | 0.06 (0.06) |
| Constant | 3.62*** (0.06) | 1.69*** (0.11) | 3.88*** (0.04) |
| N | 4,220 | 4,220 | 4,220 |
| R-squared | 0.04 | 0.10 | 0.04 |

Models 1 and 3 are linear regressions and model 2 is a logistic regression. Standard errors between parentheses.
\*\*\* $p < .001$ , \*\* $p < 0.01$ , \* $p < .05$

## DISCUSSION

This study examined the proximal effects of a mobile CDSS on physicians’ perceived clinical update, adherence to guideline-based recommendations, and confidence in clinical decision-making. Across all three domains, recent CDSS use was associated with small yet statistically significant improvements, suggesting that even brief, real-world exposure to evidence-based content may enhance both cognitive and affective dimensions of clinical reasoning.

Consistent with prior systematic reviews, our findings reinforce that CDSS contribute to improved adherence to evidence-based recommendations across diverse medical contexts— including oncology, cardiology, perioperative care, and respiratory medicine (6,12,13,17). These results align with the well-established role of CDSS in standardizing clinical practice and reducing unwarranted variation. However, beyond objective decision accuracy, our study extends current literature by addressing dimensions rarely explored in CDSS evaluations: perceived clinical update and confidence. By capturing both objective and subjective effects, this research provides a more comprehensive understanding of how digital tools shape physician behavior.

This multidimensional approach represents a conceptual advancement. In a recent systematic qualitative review, Wosny et al. (2023) found that only 17 out of 1,175 studies (1.45%) examined clinicians’ experiences with digital tools, most of which (59%) focused on CDSS. Reported experiences ranged from positive aspects—such as confidence, sense of responsibility, and satisfaction—to negative ones, including frustration, cognitive overload, and fear. The authors proposed a framework linking use, experience, and outcomes, emphasizing that clinicians’ thoughts and emotions remain underexplored and calling for research that examines experience as a mediator of professional well-being(11) . Our findings align with this perspective, suggesting that CDSS exposure may enhance clinicians’ sense of control and certainty during reasoning, bridging the gap between decision support and professional self-efficacy.

The observed effect sizes were modest, which is expected given the study’s focus on routine, low-complexity conditions. Prior literature has emphasized that CDSS are designed to complement rather than replace clinical reasoning, helping clinicians integrate multiple data sources and contextual factors to reach appropriate decisions (1). This suggests that their value may be particularly relevant in decisions requiring higher cognitive integration.

This study has limitations. The non-experimental design precludes causal inference, and the reliance on self-reported CDSS use introduces potential recall bias. Nevertheless, such misclassification would likely attenuate the associations, rendering our estimates conservative. Additionally, due to limitations in our data-collection method — which involved sending the survey up to 24 hours after the user accessed the content — our measure is also conservative because it does not assess the physician’s decision at the exact moment of consulting the CDSS. Instead, it evaluates the physician’s retention of the updated information within a 24-hour window. It is possible that the observed impact is even greater at the precise moment of decision-making, when the content is being consulted in real time. Future research with experimental design may assess the effect of accessing updated information at the exact moment of the clinical decision.

Furthermore, the analysis focused on common conditions with standardized management pathways; therefore, the observed effects may underestimate the CDSS’s impact in complex scenarios requiring broader differential diagnosis or judgment under uncertainty.

Future investigations should employ experimental or longitudinal designs to assess causal mechanisms linked to CDSS exposure, cognitive processing, and clinical outcomes. The rapid evolution of generative AI–based CDSS warrants examination of how interactivity, explainability, and personalization influence physicians’ confidence and learning. Research across diverse clinical settings and health systems, particularly in resource-limited contexts, will be essential to understand how these tools can equitably support clinical excellence.

Clinically, our findings highlight the role of digital decision-support tools not only as instruments for accuracy but also as catalysts of continuous learning and cognitive reinforcement. Theoretically, they underscore the need to integrate proximal cognitive and affective constructs—such as perceived update and confidence—into implementation science models evaluating CDSS adoption and impact. Policymakers and healthcare organizations should consider these proximal effects when designing strategies for digital transformation aimed at improving both care quality and clinician well-being.

## Data Availability

All data produced in the present study are available upon reasonable request to the authors

## FUNDING STATEMENT

The research was completely funded by Afya, the educational institution, receiving no external sponsors.

## COMPETING INTERESTS

The educational institution behind the study also owns the Afya Whitebook platform.

## DATA AVALABILITY

Data are available upon reasonable request.

## ETHICS APPROVAL

Ethics approval was obtained from Instituto de Ensino Superior Presidente Tancredo de Almeida Neves Ethics Committee (Approval number: 7.543.478).

## Notes

### Competing Interest Statement

The educational institution of the authors behind the study (Afya) also owns the Afya Whitebook platform.

### Funding Statement

The research was completely funded by Afya, the educational institution where the authors work, receiving no external sponsors.

### Author Declarations

Ethics committee/IRB of Instituto de Ensino Superior Presidente Tancredo de Almeida Neves gave ethical approval for this work

