## appendix A for "Guideline Adherence and Subjective Effects of a Mobile Clinical Decision Support System on Physicians’ Practice: A Nationwide Survey-Based Within-Subject Study"

### APPENDIX A – VIGNETTE CLINICAL CASES AND FOLLOWING QUESTIONS

#### Assessment of up-to-date knowledge on topics

Q1. Regarding the following topics, how do you assess your level of up-to-date knowledge to make clinical decisions and adopt appropriate management? (Randomized item order for response)

On a scale from 1 to 5, where 1 = "Not at all up-to-date" and 5 = "Completely up-to-date."

| | (1) Not at all up-to-date | (2) | (3) | (4) | (5) Completely up-to-date |
|---|---|---|---|---|---|
| Stroke (Cerebrovascular Accident, CVA) | | | | | |
| Asthma Exacerbation | | | | | |
| Type 2 Diabetes Mellitus (T2DM) | | | | | |
| Infectious Gastroenteritis | | | | | |
| Arterial Hypertension | | | | | |
| Acute Myocardial Infarction (AMI) | | | | | |
| Urinary Tract Infection (UTI) | | | | | |
| Bacterial Pneumonia | | | | | |

#### Clinical Cases

In the following clinical cases, imagine that you are in your daily clinical practice and have only the patient information provided as presented. Based solely on this data, make the best possible decision, even if you consider that some information may be missing or not entirely complete.

- **CASE 1: Acute Ischemic Stroke**

Clinical case description:

A 64-year-old man presenting with aphasia and hemiparesis starting 1 hour prior. NIHSS score of 14. Non-contrast head CT without hemorrhage. Blood pressure 165/95 mmHg, blood glucose 104 mg/dL.

Related questions:

Q1. Considering the time window and the clinical data, what is the most appropriate immediate therapeutic management for this patient? (Randomized answer order)

A. Administer 300 mg of acetylsalicylic acid (ASA) and monitor clinical progression

B. Initiate antihypertensive therapy to reduce blood pressure before any other intervention
C. Repeat head CT 6 hours before deciding on treatment
D. Initiate intravenous thrombolysis with alteplase after ruling out hemorrhage

✓ Correct answer: D. Initiate intravenous thrombolysis with alteplase after ruling out hemorrhage

The patient presents with an ischemic stroke within the therapeutic window (less than 4.5 hours), with significant neurological symptoms (NIHSS 14) and no contraindications. According to AHA/ASA guidelines, he is a candidate for intravenous alteplase administration immediately after exclusion of hemorrhage on CT. His blood pressure is below the threshold that contraindicates thrombolysis (185/110 mmHg) and therefore does not require prior reduction.

Q2. What is your level of confidence in the management choice above?

A. (1) Very unconfident
B. (2) Not confident
C. (3) Somewhat confident
D. (4) Confident
E. (5) Very confident

Q3. Within the first 24 hours, the patient developed decreased level of consciousness and worsening motor deficits. What is the most appropriate immediate management? (Randomized answer order)

A. Administer ASA and repeat neuroimaging after 6 hours
B. Initiate anticoagulation with heparin to prevent new ischemic events
C. Maintain clinical observation and wait for return to baseline before intervening
D. Perform an urgent head CT and discontinue any antiplatelet medication

- Correct answer: D. Perform an urgent head CT and discontinue any antiplatelet medication

  Neurological deterioration after thrombolysis should always raise suspicion for hemorrhagic transformation. The appropriate management is to obtain an emergent head CT, suspend anticoagulant and antiplatelet therapies, and initiate intensive supportive care. Use of ASA or heparin without imaging confirmation is contraindicated.

Q4. What is your level of confidence in the management choice above?

A. (1) Very unconfident
B. (2) Not confident
C. (3) Somewhat confident
D. (4) Confident
E. (5) Very confident

Q5. In the past 24 hours, have you consulted or updated yourself with decision-support information (e.g., Whitebook) regarding the topic of Stroke?

A. Yes
B. No

- **CASE 2: Moderate Asthma Exacerbation**

Clinical case description:

A 24-year-old woman with a history of asthma since childhood presents to the emergency department with dyspnea, speaking in short phrases, and using accessory muscles. Oxygen saturation: 91% on room air; heart rate: 110 beats per minute. Pulmonary auscultation reveals diffuse wheezing. Peak expiratory flow is 55% of the predicted value. She is alert, oriented, and shows no signs of exhaustion.

Related questions:

Q1. What is the most appropriate initial management for this patient? (Randomized answer order)

A. Administration of intramuscular corticosteroid alone
B. Initiation of intravenous antibiotics and hydration
C. Use of long-acting bronchodilator with immediate discharge
D. Administration of oxygen, oral corticosteroid, and short-acting inhaled bronchodilator

- Correct answer: D. Administration of oxygen, oral corticosteroid, and short-acting inhaled bronchodilator

Q2. What is your level of confidence in the management choice above?

A. (1) Very unconfident

B. (2) Not confident
C. (3) Somewhat confident
D. (4) Confident
E. (5) Very confident

Q3. After improvement of symptoms and clinical stabilization, the patient is ready for discharge. Which measure must be implemented before discharge? (Randomized answer order)

A. Initiate long-acting bronchodilator without inhaled corticosteroid
B. Prescribe empirical antibiotics to prevent respiratory infection
C. Request spirometry before releasing the patient
D. Initiate inhaled corticosteroid and provide guidance for early follow-up within 7 days

- Correct answer: D. Initiate inhaled corticosteroid and provide guidance for early follow-up within 7 days

  In patients with moderate asthma exacerbation, once clinical improvement is achieved, it is essential to initiate or optimize controller therapy, particularly inhaled corticosteroids, as recommended by GINA/SBPT guidelines. In addition, early outpatient follow-up within 7 days should be arranged to reassess disease control and treatment adherence. Empirical antibiotics or immediate spirometry are not indicated at this stage.

Q4. What is your level of confidence in the management choice above?

A. (1) Very unconfident
B. (2) Not confident
C. (3) Somewhat confident
D. (4) Confident
E. (5) Very confident

Q5. In the past 24 hours, have you consulted or updated yourself with decision-support information (e.g., Whitebook) regarding the topic of Asthma Exacerbation? (Randomized answer order)

A. Yes
B. No

- **CASE 3 — Type 2 Diabetes Mellitus (T2DM)**

Clinical case description:

A 48-year-old woman with overweight presents for a routine consultation and reports the following laboratory results: fasting plasma glucose of 140 mg/dL and glycated hemoglobin of 7.4%. She is asymptomatic, has no known comorbidities, and has no family history of severe diabetes-related complications. She is motivated to make lifestyle changes.

Related questions:

Q1. What is the most appropriate initial management for this patient? (Randomized answer order)

A. Wait three months and repeat tests before initiating treatment
B. Immediately start two glucose-lowering medications
C. Initiate fixed-dose insulin therapy
D. Start metformin and provide guidance on lifestyle modification

- Correct answer: D. Start metformin and provide guidance on lifestyle modification

Q2. What is your level of confidence in the management choice above?

A. (1) Very unconfident
B. (2) Not confident
C. (3) Somewhat confident
D. (4) Confident
E. (5) Very confident

Q3. Which test should be requested during the initial evaluation to screen for microvascular complications of diabetes? (Randomized answer order)

A. Resting electrocardiogram
B. Monthly serial glycated hemoglobin
C. Plasma C-peptide to assess pancreatic reserve
D. Urine albumin-to-creatinine ratio and serum creatinine measurement

- Correct answer: D. Urine albumin-to-creatinine ratio and serum creatinine measurement

  The initial assessment of a patient with type 2 diabetes should include evaluation of renal function (creatinine) and screening for diabetic nephropathy using the urine albumin-to-creatinine ratio. These tests are essential to identify early microvascular complications and guide the use of nephroprotective medications when necessary.

Q4. What is your level of confidence in the management choice above?

A. (1) Very unconfident
B. (2) Not confident
C. (3) Somewhat confident
D. (4) Confident
E. (5) Very confident

Q5. In the past 24 hours, have you consulted or updated yourself with decision-support information (e.g., Whitebook) regarding Type 2 Diabetes Mellitus (T2DM)? (Randomized answer order)

A. Yes
B. No

- **CASE 4: Infectious Gastroenteritis**

<u>Clinical case description:</u>

A 32-year-old man, previously healthy, presents to the emergency department with sudden-onset watery diarrhea accompanied by nausea, crampy abdominal pain, and fever of 38.2 °C, lasting 2 days. He reports more than eight bowel movements per day, with no visible blood in the stool. He reports difficulty tolerating oral fluids due to episodes of vomiting. On physical examination, he is mildly dehydrated, with dry mucous membranes and blood pressure of 100/70 mmHg. Heart rate: 102 bpm. No abdominal tenderness on deep palpation.

<u>Related questions:</u>

Q1. What is the most appropriate initial management for this patient with suspected acute infectious gastroenteritis? (Randomized answer order)

A. Administer an antiemetic and discharge immediately with oral hydration at home
B. Prescribe empirical antibiotics for enteric bacteria and discharge
C. Order a stool parasitology test and keep the patient fasting
D. Initiate intravenous hydration and observe clinical progression

- Correct answer: D. Initiate intravenous hydration and observe clinical progression

The patient presents signs of dehydration and difficulty maintaining adequate oral hydration. According to protocols such as those from WHO and national guidelines, this warrants intravenous hydration in a supervised setting, with monitoring of clinical response. Empirical antibiotics are not indicated in non-dysenteric presentations without warning signs.

Q2. What is your level of confidence in the management choice above?

A. (1) Very unconfident
B. (2) Not confident
C. (3) Somewhat confident
D. (4) Confident
E. (5) Very confident

Q3. During observation, the patient shows improvement, with no vomiting in recent hours and reduced diarrhea. What is the most appropriate management at this point? (Randomized answer order)

A. Keep the patient hospitalized for 24 hours to complete intravenous hydration despite improvement
B. Prescribe oral antibiotics and an antispasmodic before discharge
C. Request outpatient colonoscopy and discharge with 12 hours of fasting
D. Discharge with advice on a light diet, prescription for oral rehydration solution, and return if symptoms worsen

- Correct answer: D. Discharge with advice on a light diet, prescription for oral rehydration solution, and return if symptoms worsen

  With clinical improvement and recovery of tolerance to oral fluids, the patient may be discharged with dietary guidance, at-home oral rehydration, and monitoring for warning signs. Antibiotics or antispasmodics are reserved for selected cases, and colonoscopy is contraindicated in this context.

Q4. What is your level of confidence in the management choice above?

A. (1) Very unconfident
B. (2) Not confident
C. (3) Somewhat confident
D. (4) Confident
E. (5) Very confident

Q5. In the past 24 hours, have you consulted or updated yourself with decision-support information (e.g., Whitebook) regarding the topic of Infectious Gastroenteritis? (Randomized answer order)

A. Yes
B. No

- **CASE 5: Arterial Hypertension**

Clinical case description:

A 54-year-old man with no known comorbidities presents with occasional headaches. He is not taking any medication. During the consultation, his blood pressure was measured at 162/98 mmHg. After five minutes of rest, the blood pressure was measured again, with a result of 160/96 mmHg. He is in good general condition, has no abnormalities in physical examinations, and has not undergone recent blood tests or imaging.

Related questions:

Q1. What is the most appropriate management at this moment? (Randomized answer order)

A. Wait for three additional measures in future consultations before making any decision
B. Diagnose hypertension and initiate treatment with two medications
C. Diagnose hypertension and begin treatment with lifestyle modifications only, without medication
D. Request out-of-office blood pressure monitoring (home or ambulatory) to confirm the diagnosis

- Correct answer: D. Request out-of-office blood pressure monitoring (home or ambulatory) to confirm the diagnosis

  When blood pressure ranges between 140–179/90–109 mmHg and the patient show no evidence of complications or target-organ damage, the diagnosis of hypertension should be confirmed using out-of-office methods such as ambulatory blood pressure monitoring (ABPM) or home blood pressure monitoring (HBPM).

Q2. What is your level of confidence in the management choice above?

A. (1) Very unconfident
B. (2) Not confident
C. (3) Somewhat confident

D. (4) Confident
E. (5) Very confident

Q3. Once the diagnosis of mild hypertension is confirmed in a patient without additional cardiovascular risk factors, what should be the initial management? (Randomized answer order)

A. Start aspirin and cholesterol-lowering medication
B. Initiate antihypertensive medication and recommend a low-sodium diet
C. Prescribe a beta-blocker combined with a diuretic and schedule follow-up in six months
D. Recommend lifestyle modifications for three months before considering medication use

- Correct answer: D. Recommend lifestyle modifications for three months before considering medication use

  In cases of mild hypertension without increased cardiovascular risk or evidence of target-organ damage, management may begin with lifestyle modifications (diet, physical activity, weight control, smoking cessation). If no improvement occurs after three months, pharmacology therapy may be considered.

Q4. What is your level of confidence in the management choice above?

A. (1) Very unconfident
B. (2) Not confident
C. (3) Somewhat confident
D. (4) Confident
E. (5) Very confident

Q5. In the past 24 hours, have you consulted or updated yourself with decision-support information (e.g., Whitebook) regarding the topic of Arterial Hypertension? (Randomized answer order)

A. Yes
B. No

- **CASE 6: ST-Elevation Myocardial Infarction (STEMI)**

<u>Clinical case description:</u>

A 78-year-old man with hypertension, diabetes, and chronic kidney disease presents with typical chest pain for the past 3 hours. The electrocardiogram shows 2.5 mm ST-segment elevation in leads V1 to V4. Blood pressure: 145/90 mmHg. The patient is hemodynamically stable. You are on duty at an urgent care unit, and the nearest catheterization laboratory is 3 hours away by ambulance.

<u>Related questions:</u>

Q1. What is the most appropriate management for the clinical case described above? (Randomized answer order)

A. Administer ASA, clopidogrel, and transfer immediately for coronary angiography
B. Transfer immediately for coronary angiography only
C. Administer fibrinolytic therapy and transfer immediately for coronary angiography
D. Administer fibrinolytic therapy and transfer for coronary angiography within 2 to 24 hours

- Correct answer: D. Administer fibrinolytic therapy and transfer for coronary angiography within 2 to 24 hours

  In patients with ST-elevation myocardial infarction without immediate access to primary percutaneous coronary intervention (<120 minutes), the appropriate management is to perform pharmacological fibrinolysis as early as possible and, if successful, proceed with routine coronary angiography between 2 and 24 hours (pharmaco-invasive strategy), in accordance with ESC and AHA guidelines.

Q2. What is your level of confidence in the management choice above?

A. (1) Very unconfident
B. (2) Not confident
C. (3) Somewhat confident
D. (4) Confident
E. (5) Very confident

Q3. Which of the situations below would mandate primary percutaneous coronary intervention as the required initial approach? (Randomized answer order)

A. Age over 75 years
B. Chronic kidney disease
C. ST-segment elevation in the inferior wall
D. Cardiogenic shock

- Correct answer: D. Cardiogenic shock

  Primary percutaneous coronary intervention is mandatory as the initial approach in patients with contraindications to fibrinolysis or in high-risk situations such as cardiogenic shock, acute pulmonary edema, unstable arrhythmias, or progressive clinical deterioration.

Q4. What is your level of confidence in the management choice above?

A. (1) Very unconfident
B. (2) Not confident
C. (3) Somewhat confident
D. (4) Confident
E. (5) Very confident

Q5. In the past 24 hours, have you consulted or updated yourself with decision-support information (e.g., Whitebook) regarding the topic of Acute Myocardial Infarction (AMI)? (Randomized answer order)

A. Yes
B. No

- **CASE 7: Urinary Tract Infection (UTI) in Pregnant Women**

Clinical case description:

A 28-year-old woman, 23 weeks pregnant, presents with dysuria and increased urinary frequency. Afebrile. Urinalysis shows +++ leukocytes and positive nitrites. Urine culture has been collected.

Related questions:

Q1. What is the most appropriate empiric antibiotic? (Randomized answer order)

A. Ciprofloxacin
B. Levofloxacin
C. Sulfamethoxazole + Trimethoprim
D. Cephalexin

- ✓ Correct answer: D. Cephalexin

Q2. What is your level of confidence in the management choice above?

A. (1) Very unconfident
B. (2) Not confident
C. (3) Somewhat confident
D. (4) Confident
E. (5) Very confident

Q3. What is the appropriate management immediately after starting empiric treatment? (Randomized answer order)

A. Wait for the urine culture to adjust the antibiotic
B. Discharge without follow-up
C. Hospitalize for intravenous antibiotic therapy
D. Repeat urine culture after empiric treatment

- ✓ Correct answer: D. Repeat urine culture after empiric treatment

Q4. What is your level of confidence in the management choice above?

A. (1) Very unconfident
B. (2) Not confident
C. (3) Somewhat confident
D. (4) Confident
E. (5) Very confident

Q5. In the past 24 hours, have you consulted or updated yourself with decision-support information (e.g., Whitebook) regarding the topic of Urinary Tract Infection (UTI)? (Randomized answer order)

A. Yes
B. No

- **CASE 8: Bacterial Pneumonia**

Clinical case description:

A 64-year-old man, without comorbidities, presents with fever, productive cough, and chest pain for 3 days. Respiratory rate: 22 breaths/min; $SpO_2$: 94%. Chest X-ray shows consolidation in the lower right lobe. CURB-65 score = 0.

Related questions:

Q1. What is the initial therapeutic management? (Randomized answer order)

A. Clarithromycin PO for 10 days in an outpatient regimen
B. Hospitalize and start IV ceftriaxone + azithromycin
C. Levofloxacin PO with 24-hour hospital observation
D. Amoxicillin + Clavulanate PO at home

✓ Correct answer: D. Amoxicillin + Clavulanate PO at home

Q2. What is your level of confidence in the management choice above?

A. (1) Very unconfident
B. (2) Not confident
C. (3) Somewhat confident
D. (4) Confident
E. (5) Very confident

Q3. For patients treated on an outpatient basis, what is the recommended follow-up? (Randomized answer order)

A. Mandatory repeat chest X-ray in 7 days
B. Follow up only after completing the antibiotic course
C. Follow-up in 48 hours only if clinical deterioration occurs
D. Clinical review within 72 hours to reassess progression

✓ Correct answer: D. Clinical review within 72 hours to reassess progression

Q4. What is your level of confidence in the management choice above?

A. (1) Very unconfident
B. (2) Not confident
C. (3) Somewhat confident
D. (4) Confident
E. (5) Very confident

Q5. In the past 24 hours, have you consulted or updated yourself with decision-support information (e.g., Whitebook) regarding the topic of Bacterial Pneumonia? (Randomized answer order)

A. Yes
B. No
